# Prevalence, intensity and control strategies of soil-transmitted helminth infections among pre-school age children after 10 years of preventive chemotherapy in Gamo Gofa zone, Southern Ethiopia: A call for action

**DOI:** 10.1101/2020.05.14.20102277

**Authors:** Mekuria Asnakew Asfaw, Tigist Gezmu, Teklu Wegayehu, Alemayehu Bekele, Zeleke Hailemariam, Nebiyu Masresha, Teshome Gebre

**Affiliations:** Collaborative Research and Training Centre for NTDs, Arba Minch University, Ethiopia; Department of Biology, College of Natural Sciences, Arba Minch University, Ethiopia; School of Public health, College of Medicine and Health Sciences, Arba Minch University, Ethiopia; Ethiopian Public Health Institute, Addis Ababa, Ethiopia; The Task Force for Global Health, International Trachoma Initiative, Addis Ababa, Ethiopia

**Author notes:** **Corresponding author** (MA).

**Keywords:** Soil-transmitted helminths, Pre-school age children, Kato-Katz technique, Prevalence, Intensity, Control strategies of STH, Operational research

## Abstract

**Background:** Soil-transmitted helminths (STH) (*Ascaris lumbricoides, Trichuris trichiura* and hookworm) are among the most prevalent neglected tropical diseases (NTDs) in Ethiopia. Pre-school age children (PSAC) pay high morbidity toll associated with STH infections. Site specific operational evidence is lacking on prevalence, intensity and control strategies of STH among PSAC in Ethiopia. This study is, therefore, aimed to fill that missing knowledge gap.

**Methods:** We did a community-based cross-sectional study in five districts of Gamo Gofa zone; from December 2018 to January 2019. Data were collected using pre-tested questionnaire, and the Kato-Katz technique was used to diagnose parasites egg in stool. Then data were edited, coded and entered into EpiData 4.4.2, and exported to SPSS software (IBM, version 25) for analysis.

**Results:** A total of 2462 PSAC participated in this study. Overall, the prevalence of STH was 23.5% (578/2462). As*caris lumbricoides* was the most prevalent, 18.6% (457/2462), followed by *Trichuris trichiura*, 9.2% (226/2462), and hookworm, 3.1% (76/2462). The prevalence of STH in Chencha, Dita, Deremalo, Bonke and Demba Gofa districts were 33.8% (210/622), 26.4% (109/413), 21.3% (88/414), 20.6% (128/621), and 11% (43/392), respectively. Of the total, 7.4% (181/2462) PSAC were infected with two STH species. *Ascaris lumbricodes* infections had moderate intensity (15%), and the rest had low infections intensity. It is found that 68.7% of PSAC were treated with Albendazole. This study also revealed households level data as, 39.4% used water from hand-dug well, 52.5% of need to move ≥ 30minutes to collect water, 77.5% did not treat water, and 48.9% not had hand washing facility.

**Conclusion:** This study showed that a significant proportion of pre-school age children are suffering from STH infections across districts in the study area despite preventive chemotherapy distribution for more than 10 years. Further, gaps in control strategies of STH were highlighted, which calls for action.

**Author summary:** Infections with STH pose serious public health challenge among children; it causes anemia, vitamin A deficiency, stunting, malnutrition, impaired development, and intestinal obstruction. About 4 billion people are at risk for STH infection worldwide, and with over 2 billion already infected. In Ethiopia, about 81 million people are at risk for STH infection. STH can be controlled, possibly eliminated by combined interventions such as preventive chemotherapy (PC), large scale administration of anthelmintic drugs to at-risk population, and improved water, sanitation and hygiene (WASH). Ethiopia has been working to reduce the prevalence of moderate and heavy infections with soil transmitted helminth in pre-school and school aged children to below 1%. Since 2013 remarkable progress has been made on STH control strategies in terms of mapping and distribution of PC. However, evidence is lacking to track progress towards control and elimination goal, particularly among PSAC. The findings of our study highlight that achieving STH control and elimination goal in Gamo Gofa zone by 2020 and even beyond could be challenging unless the current STH control strategies are improved.

## Introduction

Soil-transmitted helminth (*Ascaris lumbricoides, Trichuris trichiura* and hookworm) infections are among the most common neglected tropical diseases (NTDs) [1]. It is prevalent mainly in tropical and subtropical areas where water supply, hygiene and sanitation infrastructure are inadequate [2–4] . Moderate and heavy intensity of STH infections are associated with chronic harmful effects on vitamin A and iron status, physical, intellectual, and cognitive development in pre-school age children (PSAC), these morbidities not only take a huge toll on the health of children, but has also been shown to affect economic development of a nation [5, 6].

Globally, over 2 billion people are affected with STH where ascariasis accounts for almost 1.2 billion infections while trichiurasis and hookworm (*Ancylostoma duodenale* and *Necator americanus*) responsible for over 800 million and 740 million infections, respectively [1, 7, 8] . The global burden of STH infection is estimated at between 5 and 39 million disability-adjusted life years (DALYs) and in 2010, 5.18 DALYs were estimated as associated with STH infections [9, 10]. The greater burden of STH infection is found in the tropical countries including tropical South America, China, East Asia, and Sub-Saharan Africa [5]. According to the WHO estimate 42 countries in Africa are endemic for STH with 284 million cases occurring in both school age and pre-school age children. These children require periodic administration of preventive chemotherapy [1, 11].

STH are widely prevalent parasites in Ethiopia, and exceeds over 85% prevalence in some districts. The number of people living in STH endemic areas is estimated at 81 million, of which pre-school age children account 9.1 million [12]. Pre-school age children (PSAC) and school-age children (SAC) are the most exposed groups and who pay the highest toll of morbidity associated with STH infections. Significant proportions of pre-school age children are affected by STH, which account 10% to 20% of the 3.5 billion people living in STH-endemic areas. The greatest numbers of intestinal worms harbored in these children resulting in diarrhea, loss of appetite, weight loss, growth retardation, malnutrition, anemia & cognitive defects [6, 13–16].

In 2012, the World health organization (WHO) endorsed road map to combat NTDs by 2020, and substantial progress has been made in terms of controlling STH morbidity [3]. The WHO goal is to reduce the prevalence of moderate and heavy infections with soil transmitted helminths in preschool and school aged children to below 1%, to be no longer considered public health problems, by 2020 [17]. In line with this, Ethiopia has also set a similar goal to achieve by the same year [12]. To achieve these goals, in areas where prevalence of any soil-transmitted infection is 20% or higher, periodic mass administration of preventive chemotherapy (deworming) using annual or biannual single-dose Albendazole or Mebendazole is recommended by WHO for all pre-school and school age children (10). As part of the global action towards Universal Health Coverage (UHC), ending NTDs is prioritized by 2030 in the Sustainable Development Goal (SDG) under target 3.3, to ensure healthy lives and well-being for all ages groups [3]. Therefore, SDGs could be attained only if the NTD goals are met. Moreover, working on NTDs helps the vision of universal health coverage, which means that all individuals and communities access the health services they need without suffering from financial suffering [18].

Since 2008 preventive chemotherapy (PC) against STH in PSAC has been implemented alongside Vitamin A distribution in the study area as well as at national level through community based drug distribution platforms [12]. In 2013, the first national master plan for NTDs was launched, and then the government of Ethiopia has been collaborating with the WHO and other partners for mapping all endemic districts to address SAC through the school based mass drug administration [19]. This implies that PC was started before ten years to combat STH in PSAC at study area. Targeting on SAC earlier studies have shown the impact of deworming on prevalence and intensity of STH infections. On the other hand, the impact of deworming on STH infection status among PSAC is not well monitored and evaluated. Hence, evidence is lacking at national level on prevalence, intensity and control strategies of STH infections among PSAC. This condition is challenging for planning of deworming programs on STH. Furthermore, timely and reliable operational research evidence is needed to inform decision-making, and also data could be used for further actions to improve control strategies of soil-transmitted helminths among pre-school-age children. Therefore, the present study is aimed to determine prevalence, intensity and control strategies of STH infections among preschool children in Gamo Gofa zone.

## Methods

### Study area and period

This study was conducted in the former Gamo Gofa zone; from December 2018 to January 2019. The zone is found in Southern Nations, Nationalities, and Peoples’ Regional State (SNNPR) and it had 15 districts and two city administrations (CAs). All districts and CAs are endemic for STH, 15 had moderate prevalence and two had low prevalence According to the 2007 estimate of Central Statistical Agency of Ethiopia, a total of 2,043,668 people live in the zone, of which 1,013,533 are males and 1,030,135 are females [20]. Arba Minch is the capital city of the zone, which is located at 437 km away from Addis Ababa, capital city of Ethiopia.

### Study design and populations

Community-based cross-sectional study was conducted to determine prevalence, intensity and control strategies of STH infections among PSAC in the study area. The study population was all PSAC (1–5 years) in the selected STH endemic *Kebeles* (localities). PSAC who were unable to give stool samples at the time of collection were excluded from the study. Additionally, PSAC were excluded in case when they were seriously ill or parents were unable to provide their information.

### Determining sample size and sampling technique

The sample size was determined using single population proportion formula,

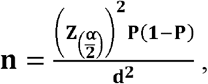

assuming p, **25.7%** (proportion of PSAC infected with STH) [21]; **Z**, 1.96 at significant level of alpha **(α)** of **0.05**, and desired degree of precision (d) of **3 %**, and design effect = **3**. The computed sample size was **2434** and by adding **10%** non-response rate, the total computed sample size was **2678**.

Multi-stage cluster sampling technique was employed in order to select study participants (Fig 1). First, districts and CAs STH endemicity status was identified based on results of previous STH mapping survey conducted at national level (where 2 had low and 15 had moderate prevalence level [22]. We excluded 2 districts with low endemicity status since they were not eligible for preventive chemotherapy. Two, 5 districts and 12 *kebeles* were selected from 15 districts by simple random sampling technique (SRS). Three, list of eligible households in each *kebeles* which had children between 1 and 5 years of age were identified by health extension workers (HEWs). Finally, children from each *kebeles* were selected by taking probability proportionate to sample size into account.

**Figure 1.**
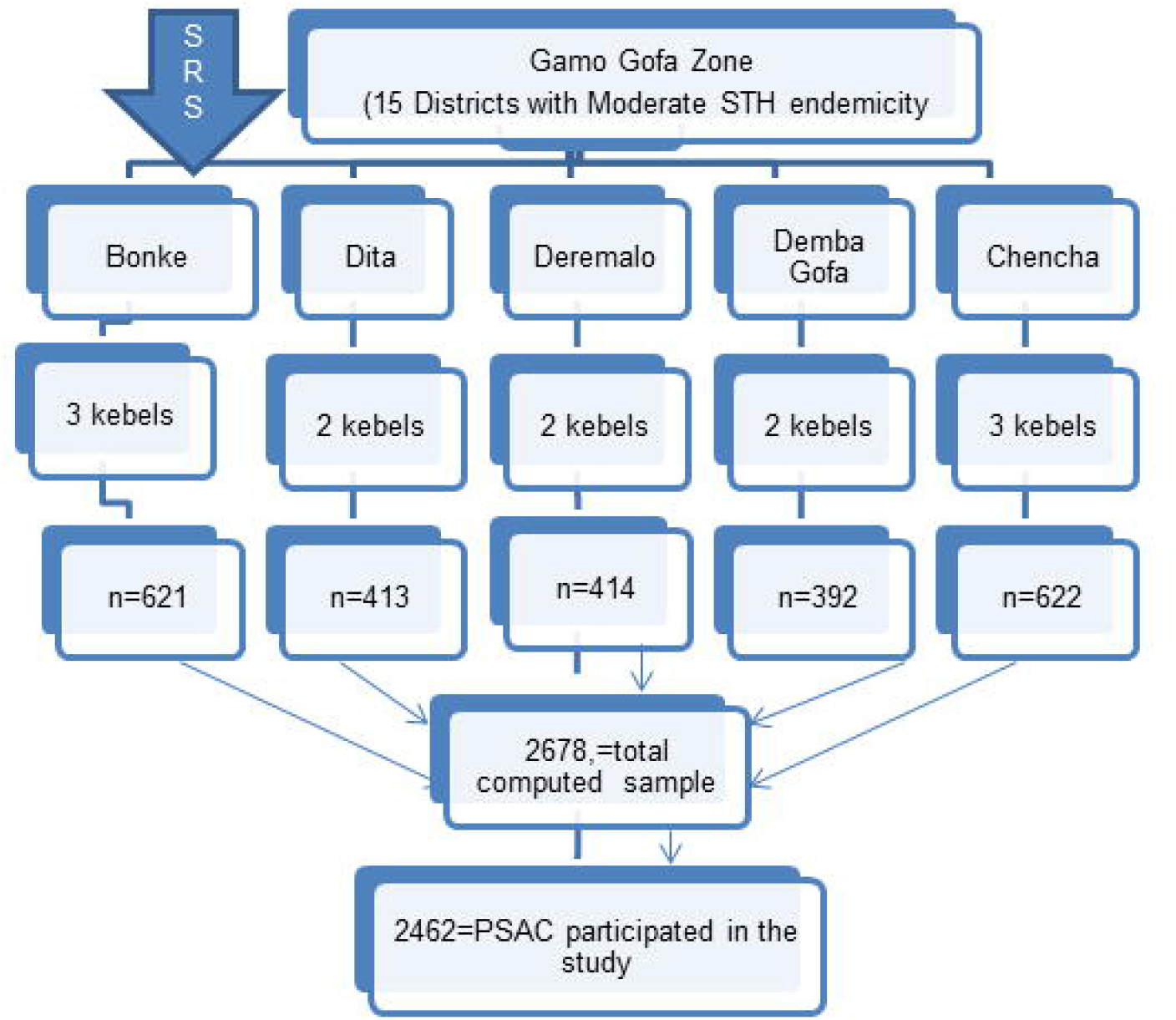
Sampling profile

### Study variables and data collection

Variables included in this study were STH infection status (positive or negative), intensity of infections, socio-demographic characteristics of parent and guardians, social determinants of health, wealth status of household and child related variables and implementation of STH control strategies. Data on these socio-demographic and other variables were collected through face-to-face interviews using pre-tested questionnaire with heads of households or mothers or guardians using pre-tested questionnaire. Stool specimens were examined using the WHO recommended Kato-Katz kit under microscopy [23].

### Stool collection and processing

Fresh stool specimens were collected using clean, leak proof and screw cup container, and then collected stool samples were transported in cool boxes to nearby health facility for examination. The stool samples were processed within two hours of receipt or saved in Icebox where travel time exceeded two hours. Samples were examined in the local health center by Kato-Katz technique to determine the prevalence and intensity of STH. Duplicate slides were prepared per stool sample in order to ensure reliability.

### Quality assurance

Data quality was monitored through standard operational procedure (SOPs), recruiting competent data collectors, pre-testing tools, training data collectors and supervisors, daily checking consistency and accuracy of collected data. The quality of data collection was closely monitored by supervisors. Stools were examined by a qualified laboratory technologist. For the purpose of bench aid, pictures of parasites egg were displayed on wall in front of microscopy examination for the purpose of internal quality control.

### Statistical data analysis and measurements

First, data were edited, coded and entered into EpiData 4.4.2, and then exported to SPSS software (IBM, version 25) for analysis. A difference in prevalence of STH between or among categories of variables was analyzed using chi-square test (X^2^). Summary statistics were computed, and data were presented with tables and graphs using excel spreadsheet. Children between age 1 and 5 years were defined as PSAC [23]. Wealth analysis was performed as, initially, reliability test was performed using the economic variables involved in measuring the wealth of the households. The variables which were used to compute the alpha value were entered into the principal component analysis. At the end of the principal component analysis, the wealth index was obtained as a continuous scale of relative wealth. Then, quintiles of the wealth index were created.

We calculated the prevalence by dividing the number of STH positive PSAC by the total number of participants. Intensity of STH infection is the number of helminths (worms) infecting an individual; for each parasite species it was analyzed as light, moderate and heavy infections based on eggs per gram of stool (EPG) and was classified according to the WHO guidelines [24], as follow (Table 1).

**Table 1.** Criteria for classifying intensity of STH infection for each species. STH infection Intensity of infection (EPG)

| STH infection | Intensity of infection (EPG) |  |  |
| --- | --- | --- | --- |
|  | Light | Moderate | Heavy |
| *A.lumbricoides* | (1–4999) | (5000–49999) | (≥50000) |
| *T.trichiura* | (1–999) | (1000–9999) | (≥10000) |
| *Hookworms* | (1–1999) | (2000–3999) | (≥4000) |

According to WHO STH endemicity mapping classifications, there are three categories in line with implementation of mass drug administration (MDA): i) high transmission (where prevalence is > 50%), ii) moderate transmission (where prevalence is between 20%- 50%), and iii) low transmission (where prevalence is < 20%) [23, 25].

### Ethical considerations

Ethical approval was obtained from Institutional and Research Ethics review board of Arba Minch University, College of Medicine and Health Sciences, Arba Minch, Ethiopia. Oral and written consent was sought from district administrators and heads of the household before survey was conducted. Children tested positive for one or more STH were treated with Albendazole or Mebendazole by health workers.

## Results

### Socio-demographic characteristics of PSAC and heads of household

A total of 2462 (92%) PSAC participated in this study. Of the total, 246 (10%) were under 2 years of age, and slightly more males participated than females (52% versus 48%). The average number of people living in a household was 5.5 (standard deviation ±1.8) individuals. More than half of HHs (57.5%) did not attend any formal education (Table 2).

**Table 2.** Socio-demographic characteristics of PSAC and HHs, 2019 (N = 2462)

| Variables | Category | Frequency | % |
| --- | --- | --- | --- |
| Child sex | Male | 1281 | 52 |
|  | Female | 1181 | 48 |
| Child age (years) | Less than 2yrs | 246 | 10 |
|  | 2-5yrs | 2216 | 90 |
| Age of HH (years) | Less than 20 | 22 | 0.89 |
|  | 20 -29 | 404 | 16.41 |
|  | 30-39 | 1284 | 52.15 |
|  | 40-49 | 647 | 26.28 |
|  | 55-59 | 59 | 2.4 |
|  | 60 and above | 46 | 1.87 |
| Sex of HH | Male | 2174 | 88.3 |
|  | Female | 288 | 11.7 |
| Educational status | Can't read and write | 1073 | 43.58 |
|  | Can read and write | 343 | 13.93 |
|  | Elementary | 508 | 20.63 |
|  | Secondary | 279 | 11.33 |
|  | Diploma and above | 259 | 10.52 |
| Occupation | Farming | 1606 | 65.23 |
|  | Employed | 312 | 12.67 |
|  | Merchant | 303 | 12.31 |
|  | Unemployed | 78 | 3.17 |
|  | Others* | 163 | 6.62 |
| Family size | Less than 4 | 288 | 11.7 |
|  | 4-6 | 1503 | 61.05 |
|  | 7 and more | 671 | 27.25 |
| Residence | Urban | 628 | 25.5 |
|  | Rural | 1834 | 74.5 |
| Wealth quintile | Wealthiest | 488 | 19.82 |
|  | Wealthy | 498 | 20.22 |
|  | Middle income | 482 | 19.58 |
|  | Poor | 498 | 20.23 |
|  | Poorest | 496 | 20.15 |
| Water source | Pipe | 1128 | 45.82 |
|  | Well | 293 | 11.9 |
|  | Public bono | 971 | 39.44 |
|  | Other** | 70 | 2.84 |

\*=Daily laborer; \*\*=river, well and spring

### Prevalence of STH infections

Of the children surveyed, 23.5% (578/2462) had at least one type of STH infection. Ascariasis was the most prevalent (18.6%), followed by trichiurasis (9.2%) and hookworms(3.1%). Mixed STH infections (*Ascaris lumbricoides* and *Trichuris trichiura*) were found in 7.4% of PSAC. The highest prevalence of any one of STH infection was observed in Chencha district (33.8 %), as contrasted to the lowest prevalence that was found in Demba Gofa district (11%). In Deremalo district, considerable amount of hookworm infections (10%) were revealed (**Fig 2**).

**Fig 2.**
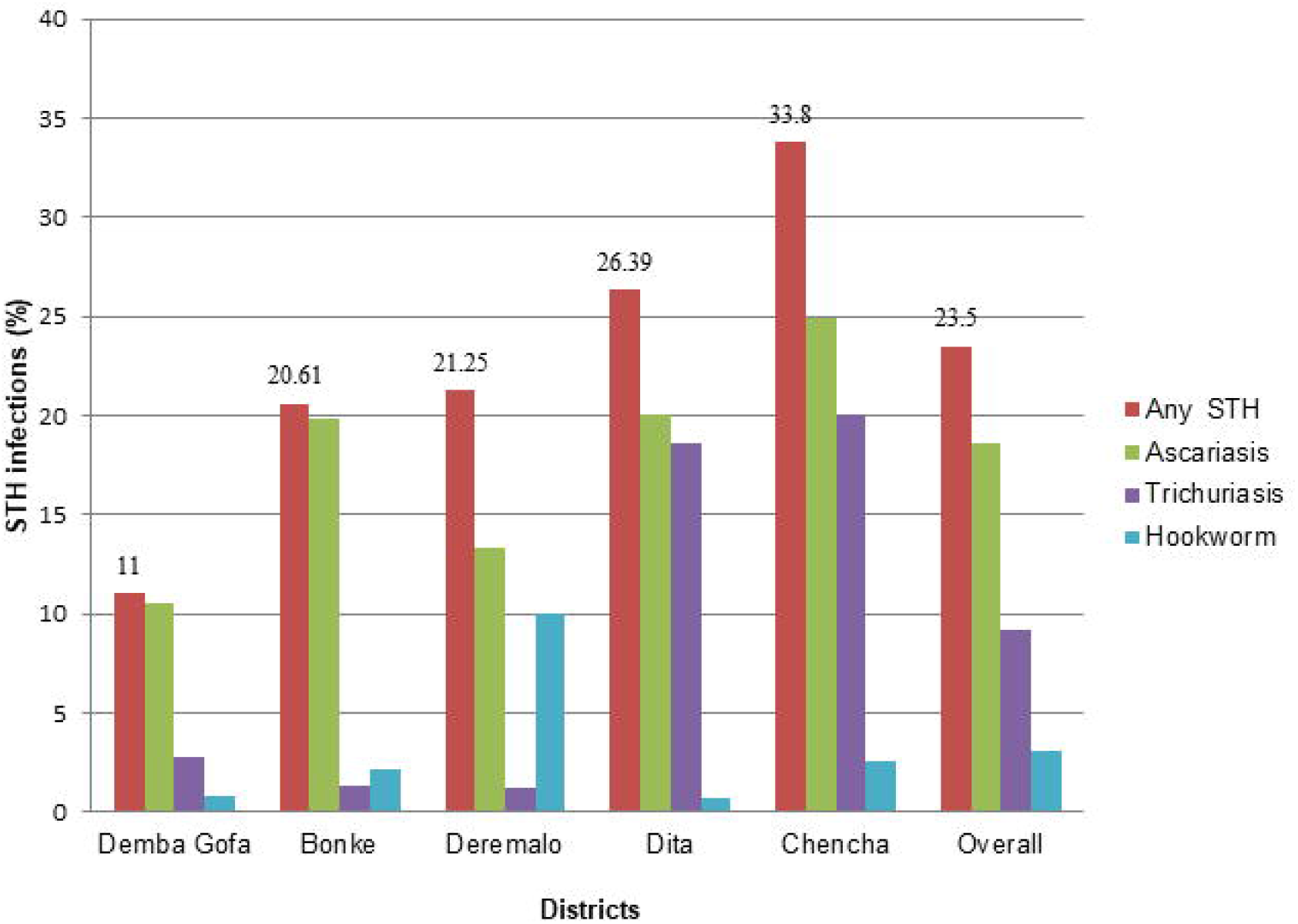
Prevalence of any STH infection by district among PSAC, Gamo Gofa zone, Southern Ethiopia, 2019 (N = 2462).

The prevalence of STH infections across the age-group (1 – 5 years) was slightly higher at ≤ years while comparing across the age-group (1–5 years). On the other hand, prevalence of any STH infections among females (24.3%) was a little higher than the male’s (22.7%), and a higher prevalence of STH infections (24.6%) in rural area was noticed than urban (20.1%). Children who did not receive PC in the last one year had slightly higher prevalence of STH infections compared to those who received PC (23.9% versus 18.7%), (Table 3).

**Table 3.** Prevalence of STH infections in association with different factors among PSAC in Gamo Gofa zone, 2019 (N = 2462).

| Variable | Category | STH Infection status |  | X2-test | P-value |
| --- | --- | --- | --- | --- | --- |
|  |  | Negative, n (%) | Positive, n (%) |  |  |
| **Child sex** | Male | 990(77.3) | 291(22.7) | 0.354 | 0.366 |
|  | Female | 894(75.7) | 287(24.3) |  |  |
| **Child age group (years)** | Less than 2yrs | 187(76) | 59(24) | 0.039 | 0.843 |
|  | 2-5yrs | 1697(76.6) | 519(23.4) |  |  |
| **Place of residence** | Urban | 502(79.9) | 126(20.1) | **5.467** | **0.019** |
|  | Rural | 1382(75.4) | 452(24.6) |  |  |
| **Child age (years)** | 1 | 187(76) | 59(24) | 0.914 | 0.923 |
|  | 2 | 414(75.1) | 137(24.9) |  |  |
|  | 3 | 563(77) | 168(23) |  |  |
|  | 4 | 522(77.2) | 154(22.8) |  |  |
|  | 5 | 198(76.7) | 60(23.3) |  |  |
| **Dewormed in the last one year** | No | 608(78.9) | 163(21.1) | 3.408 | 0.065 |
|  | Yes | 1276(75.5) | 415(24.5) |  |  |
| **Child soil eating habit** | No | 1518(76.9) | 457(23.1) | 0.371 | 0.542 |
|  | Yes | 261(78.4) | 72(21.6) |  |  |
| **Family size** | <4 | 220(76.4) | 68(23.6) | 2.219 | 0.330 |
|  | 4-6 | 1164(77.4) | 339(22.6) |  |  |
|  | 7 and above | 500(74.5) | 171(25.5) |  |  |
| **Mothers' (guardians') educational status** | Can't read and write | 774(76.9) | 233(23.1) | 5.432 | 0.246 |
|  | Can read and write | 291(73.9) | 103(26.1) |  |  |
|  | Elementary | 486(76.1) | 153(23.9) |  |  |
|  | Secondary | 225(76.8) | 68(23.2) |  |  |
|  | Diploma and above | 108(83.7) | 21(16.3) |  |  |
| **Mothers' (guardians') occupation** | Farming | 974(74.6) | 331(25.4) | **16.288** | **0.003** |
|  | Employed (Gov.) | 166(81.8) | 37(18.2) |  |  |
|  | Merchant | 348(73.7) | 124(26.3) |  |  |
|  | Unemployed | 334(82.3) | 72(17.7) |  |  |
|  | Others* | 62(81.6) | 14(18.4) |  |  |
| **Wealth quintile** | Wealthiest | 379(77.7) | 109(22.3) | 1.372 | 0.712 |
|  | Wealthy | 379(76.1) | 119(23.9) |  |  |
|  | Middle income | 374(77.6) | 108(22.4) |  |  |
|  | Poor | 369(74.1) | 129(25.9) |  |  |
|  | Poorest | 383(77.2) | 113(22.8) |  |  |
| **Water source** | Pipe | 891(79) | 237(21) | **8.289** | **0.04** |
|  | Well | 223(76.1) | 70(23.9) |  |  |
|  | Public bono | 721(74.3) | 250(25.7) |  |  |
|  | Other** | 49(70) | 21(30) |  |  |

\*=Daily laborer and housewife
\*\*=River, well and spring

### Intensity of STH infections

In majority of STH infections (85%) low infection intensity was found as a result of *hookworms* and *Trichuris trichiura* infection while 15% of Ascariasis had moderate infection intensity (Table 4). All of the moderate infections were from Chencha and Bonke district.

**Table 4.** Intensity of STH infection among PSAC in Gamo Gofa zone, southern Ethiopia, 2019 (N = 2462).

| Type of STH infection | Mean (EPG) | Intensity of infection |  | Total infected PSAC |
| --- | --- | --- | --- | --- |
|  |  | Light | Moderate |  |
| Ascariasis | 2152 | 388(84.90) | 69(15.1) | 457 |
| Hookworm | 154 | 76(100) | 0 | 76 |
| Trichiurasis | 135 | 226(100) | 0 | 226 |

### Control strategies of STH

### Treatment coverage of Albendazole

The treatment coverage of Albendazole (ALB) in the last one year before the survey against STH among PSAC was 68.7% (1691/2462) (**Fig 3**).

**Figure 3.**
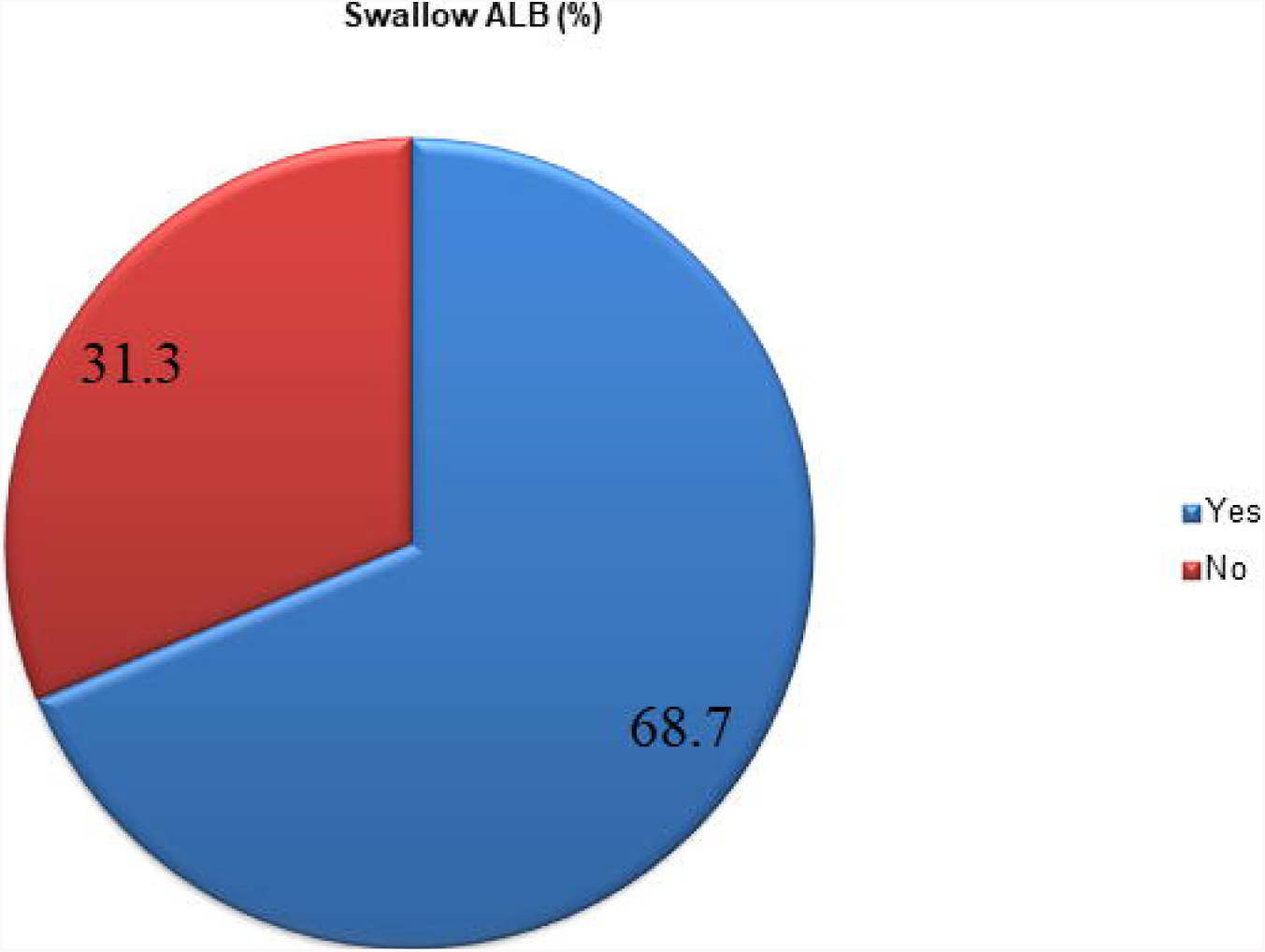
Treatment coverage of Albendazole against STH in Gamo-Gofa Zone, 2019 (N = 2462).

### Knowledge and practice (KP) of mothers or guardians on STH

Of the total surveyed mothers or guardians, the mean KP score (≤5) on transmission of STH was 92% (2262/2462); 42.7% (1052/2462) did not wash hand after defecation, and 77.7% (1913/2462) got information on STH from health extension workers (HEWs) (Table 5).

**Table 5.** Knowledge and practice on STH transmission and prevention among mothers, 2019 (N=2462).

| **Variables** | **Category** | **Frequency** | **%** |
| --- | --- | --- | --- |
| **Know about STH** | Yes | 2208 | 89.7 |
|  | No | 254 | 10.3 |
| **Know at least one STH transmission (n=2208)** | Yes | 2153 | 97.5 |
|  | No | 55 | 2.5 |
| **Know at least one STH prevention way (n=2208)** | Yes | 2135 | 96.7 |
|  | No | 73 | 13.5 |
| **Mean KP score on transmission STH** | ≤5 | 2262 | 91.9 |
|  | >5 | 200 | 8.1 |
| **Mean KP score on transmission STH** | ≤5 | 2283 | 92.7 |
|  | >5 | 179 | 7.3 |
| **Get information about STH through** | Health facility | 271 | 11 |
|  | HEWs | 1913 | 77.7 |
|  | Radio or TV | 24 | 1 |
|  | None | 254 | 10.3 |
| **Wash hand after latrine** | Yes | 1410 | 57.3 |
|  | No | 1052 | 42.7 |
| **Wash hand before meal** | Yes | 2347 | 95.3 |
|  | No | 2462 | 4.7 |
| **Wash hand after cleaning child** | Yes | 2229 | 90.5 |
|  | No | 233 | 9.5 |
| **Wash hand before cooking** | Yes | 2069 | 84 |
|  | No | 393 | 16 |
| **Wash fruit before eating** | Yes | 1841 | 74.8 |
|  | No | 621 | 25.2 |
| **Wash hand after work** | Yes | 2253 | 91.5 |
|  | No | 209 | 8.5 |

### Water, Sanitation and Hygiene (WASH)

This study also revealed households data as, 39.4% used water from hand-dug well, 52.5% of need to move ≥30minutes to collect water, 77.5% did not use treat water, and 48.9% did not own hand washing facility (Table 6).

**Table 6.** WASH characteristics of households in Gamo Gofa zone, 2019.

| Variables | Category | Frequency | % |
| --- | --- | --- | --- |
| Source of water | Pipe | 1128 | 45.82 |
|  | Well | 293 | 11.9 |
|  | Public bono | 971 | 39.44 |
|  | Other* | 70 | 2.84 |
| Distance form water source | <30min | 1170 | 47.5 |
| | $\geq 30$min | 1292 | 52.5 |
| Adequate water | No | 536 | 21.8 |
|  | Yes | 1926 | 78.2 |
| Ever treat water | No | 1909 | 77.5 |
|  | Yes | 553 | 22.5 |
| Latrine | Yes | 2397 | 97.4 |
|  | No | 65 | 2.6 |
| Type of latrine (n=2397) | Pit | 2360 | 98.5 |
|  | Improved pit latrine | 37 | 1.5 |
| Feces observed inside the latrine compound | No | 1766 | 73.67 |
|  | Yes | 631 | 26.32 |
| Hand washing facility (functional) | No | 1225 | 51.1 |
|  | Yes | 1172 | 48.9 |
| Soap or ash available (n=1172) | Yes | 298 | 25.4 |
|  | No | 874 | 74.6 |
| Reason not latrine available (n=65) | No space | 6 | 9.2 |
|  | No money | 10 | 15.4 |
|  | No skill | 9 | 13.8 |
|  | Did not know importance | 40 | 61.5 |
| If no latrine where you defecate (n=65) | Open field | 56 | 86.2 |
|  | Public | 9 | 13.8 |

\*=river, well and spring

## Discussion

This study provides operational evidence on prevalence, intensity and control strategies of soil-transmitted helminth infections (STH) to improve control strategies of soil-transmitted heliminths in Gamo Gofa zone. Our study showed that a significant proportion of pre-school age children are suffering from STH infections in the study area despite preventive chemotherapy distribution for more than 10 years. Considerable variation in prevalence, and intensity of infections across districts were also noticed. In addition, gaps in the control strategies of STH to control and eliminate STH were highlighted.

In this study, the overall prevalence of STH infections with at least one STH parasite was 23.5%, would be classified into the moderate transmission category (where prevalence is between 20% and 50%), and qualifies the requirement of annual STH mass drug administration [17]. Ascariasis was the most prevalent infection (18.6%), followed by trichiurasis (9.2%) and hookworm (3.1%). All infections of *hookworm* and *Trichuris trichiura* had low intensity while Ascariasis had moderate intensity (15 %). Despite initiation of preventive chemotherapy about 10 years ago, the burden of STH did not show significant reduction probably due to weak implementation of control strategies such as social behavioral change communication (SBCC), inadequate mass drug administration coverage and WASH as highlighted in the findings of this study.

The overall prevalence of STH infections observed in this study is comparable with studies conducted in another parts of Ethiopia (Butajira and Wonji) and West China [26–28], but slightly lower than the prevalence reported in Dembiya, northwest Ethiopia [21]. In addition, the prevalence in our study is significantly lower than the findings of other studies conducted in another part of Nigeria, Cameroon, Ecuador, Uganda, Kenya and Honduras [29–36]. These differences observed from our study could be due to variation in socio-cultural, social determinants, behavioral characteristics and implementation of prevention and control measures.

In this study, Ascariasis was identified as the commonest species of STH, and this findings supports studies done in other part of Ethiopia, Nigeria and China [21, 26, 28, 29]. On the contrast, some other studies conducted in Ethiopia, Ecuador and Honduras showed high prevalence of Trichiurasis [32, 35, 37], and a study conducted in Uganda showed high prevalence of hookworm [33]. These differences might be related to variation in environmental factors such as climate, rainfall, topography, surface temperature, altitude, and soil type which have a great impact on the distribution of STH [38]. Moreover, in this study we found significant amounts of mixed infections (7.4%) of PSAC were infected with two STH species (*Ascaris lumbricodes and Trichuris trichiura*); this finding is in line with a study conducted in other areas of Ethiopia and Nigeria [26, 27, 29].

In our study, slightly higher prevalence at age ≤2 years was observed while comparing across the age-group (1–5 years); the possible explanation related to this difference is due to the fact that current mass drug administration among PSAC often do not include age ≤2 years. On the contrast, other studies revealed numerical increase in prevalence of STH as age increase [26, 29]. In addition, in this study, prevalence of any STH infection among females (24.3%) was a little higher than the male’s prevalence (22.7%), this result supports the findings of a study conducted in another part of Ethiopia [26]. These differences might be due to variation in socio-cultural pattern, access and uptake of preventive chemotherapy.

In addition, significant proportion (15%) of moderate intensity Ascariasis was observed in our study, and this finding is higher than results of a study conducted in Butajira, Ethiopia and Honduras [26, 35]. The possible explanation related to this difference could be consistency and frequency of mass drug administration that may affect intensity of infections [23].

Furthermore, findings of our study showed inadequate implementation of control strategies. The treatment coverage of Albendazole (ALB) in this survey against STH among PSAC was (68.7%); which is lower than the national coverage (71%) of Ethiopia and WHO’s target (minimum of 75%) [10, 39]. Most importantly, study participants in this study were pre-school age children, who might not be reached out by the deworming program, especially those under 2 years old. On the other hand, obviously, school age children could have better chance to be reached by school-based deworming.

The importance of WASH intervention to control and eliminate STH was reported in different studies though site-specific data are required in our case [40–42]. However, in this study inadequate WASH infrastructures were observed at household level where 39.4% were using water from well, 52.5% were walking ≥30minutes to collect water, 77.5 % did not treat water, and 48.9% of not had hand washing facility. Of the total surveyed mothers or guardians, the mean KP score (≤5) on transmission of STH was 92% (2262/2462); 42.7% (1052/2462) did not wash hand after defecation, and 77.7% (1913/2462) got information on STH from health extension workers (HEWs). The possible reason for these findings could be related to weak social behavioral change communication (SBCC) intervention.

The most outstanding strength of our study is that it is addressing an important national operational research priority which is focusing on parasitological monitoring and control strategies of STH among pre-school age children.

In this study, there are limitations that need to be taken into account. There might be underestimation of prevalence of STH due to the fact that [1] we collected single stool specimen, which could cause variation in egg excretion over different times (hours) within a day and across different days; [2] samples were collected from remote villages and there might be rapid desiccation of hookworm eggs in the stool samples, and [3] even though the Kato–Katz technique is sensitive in detecting moderate and high infection intensity (MHI), it has lower detection power and therefore lower positive predictive values in low-prevalence (and low-intensity) settings [28, 43].

## Conclusion

Data from our study showed that substantive proportion of pre-school age children in the study area are suffering from significant burden of STH infections despite provision of preventive chemotherapy distribution for more than 10 years. Considerable variation in prevalence and intensity of infections across districts and *kebeles* was noticed. Moreover, the study highlights the need for implementing control strategies of STH in a coordinated and regular manner in order to achieve the national goal of STH control and elimination. Further, operational research needs to be conducted in different settings to monitor and evaluate progress towards elimination and control of STH among PSAC.

## Data Availability

All data are fully available without restriction

## Acknowledgements

Authors would like to thank study participants, data collectors, supervisors, zonal health office heads, district health office heads, NTDs focal persons in all selected districts of the study area and Ethiopian public health institute for their support in the implementation of the study.

## Author’s contributions

MA prepared the original draft. MA, TG, TW, AB, ZH, and TG^5^ involved in conception, protocol development and design of the study. MA, TW and TG involved in supervision of the project. MA and NM performed data cleaning and analysis. All authors were involved in the interpretation of the data. MA, TW and TG^5^ critically reviewed the initial manuscript. All authors reviewed and approved the final manuscript.

